# Heterologous Gam-COVID-Vac (Sputnik V) / mRNA-1273 (Moderna) vaccination induces a stronger humoral response than homologous Sputnik V in a real-world data analysis

**DOI:** 10.1101/2022.04.08.22273532

**Authors:** Matías J. Pereson, Lucas Amaya, Karin Neukam, Patricia Bare, Natalia Echegoyen, María Noel Badano, Alicia Lucero, Antonella Martelli, Gabriel H. Garcia, Cristina Videla, Alfredo P. Martínez, Federico A. Di Lello

## Abstract

**Introduction:** Growing data are demonstrating safety and immunogenicity of heterologous vaccination schemes against severe acute respiratory syndrome coronavirus-2 (SARS-CoV-2) infection. This strategy opens up the possibility of a shorter path towards the end of the pandemic.

**Objective:** To compare the homologous prime-boost vaccination scheme of Gam-COVID-Vac (Sputnik V, SpV) to its heterologous combination with mRNA-1273 (Moderna, Mod) vaccine.

**Methods:** SARS-CoV-2 anti-spike (S)-receptor binding domain (RBD) IgG concentration was assessed three to seven weeks after complete vaccination. Reactogenicity was evaluated by declared side events and medical assistance required until day 7 post-boost.

**Results:** Of 190 participants enrolled, 105 received homologous SpV/SpV and the remaining heterologous SpV/Mod vaccination scheme, respectively. Median (interquartile range, IQR) age was 54 (37-63) years, 132 (69.5%) were female and 46 (24.2%) individuals had a prior confirmed COVID-19. Anti-S-RBD IgG median (IQR) titers were significantly higher for SpV/Mod [2511 (1476-3992) BAU/mL] than for SpV/SpV [582 (209-1609) BAU/mL, p<0.001] vaccination scheme. In a linear model adjusted for age, gender, time to the serological assay and time between doses, SpV/Mod [4.154 (6.585-615.554), p<0.001] and prior COVID [3.732 (8.641-202.010), p<0.001] were independently associated with higher anti-S-RBD IgG values. A higher frequency of mild-moderate adverse effects was associated with the heterologous scheme, although it was well tolerated by all individuals and no medical assistance was required.

**Conclusion:** The heterologous SpV/Mod combination against SARS-CoV-2 is well tolerated and significantly increases humoral immune response as compared to the homologous SpV/SpV immunization.

## INTRODUCTION

Vaccination is the best strategy to limit the pandemic caused by the severe acute respiratory syndrome coronavirus-2 (SARS-CoV-2). The heterologous vaccination schemes approval represents a valuable alternative to the fluctuating supply of the second doses of certain homologous schemes and reliefs concerns about the side effects of heterologous schemes in people at high risk of serious adverse effect^1,2^. Studies on the heterologous vaccination schemes including the adenovirus vaccine ChAdOx1 (Vaxzevria, AstraZeneca, Oxford, UK) followed by the mRNA vaccines Pfizer (BNT162b2 Comirnaty, BioNTech, Mainz, Germany) or the mRNA-1273 (Moderna, Cambridge, MA, USA) have yielded encouraging results. In this sense, robust humoral and cellular immune response, safety, and enhanced neutralizing activity have been proved for these heterologous schemes^3-8^.

Gam-COVID-Vac (Sputnik V) is a Russian recombinant adenovirus-based vaccine (adenovirus 26 for prime and adenovirus 5 for boost) that has been approved for emergence use in more than 71 countries including Argentina^9^. Sputnik V has proven safety and efficacy (91.6%) in phase 2/3 clinical trials and effectiveness after the first (78.6-87.6%) and two components (75.5-100%) administration in real-life studies^10-13^. The Gamaleya Research Centre recommended a 21-day interval between the first and the second dose, but they also stated that it is possible to increase the interval from the earlier approved 21 days to up to three months. Interval extension does not affect the vaccine-induced immune response^14^.

In Argentina, the Sputnik V second component delayed supply has led to the implementation of a heterologous vaccination scheme with the Moderna vaccine. However, there is no information about the heterologous Sputnik V/Moderna immune response.

Humoral immune response to vaccination represents the most used tool to evaluate vaccine performance; in addition, it has been correlated with protective effects against symptomatic SARS-CoV-2 infection^15^. The aim of this study was to determine the immunogenicity and reactogenicity of the Moderna vaccine administered as boost to individuals primed with Sputnik V

## MATERIAL AND METHODS

### Study design and population

In this observational cohort study, all subjects who attended to the Centro de Educación Médica e Investigaciones Clínicas “Norberto Quirno” (CEMIC), Buenos Aires, Argentina, from December 2020 to August 2021 were included if i) they had received Sputnik V prime immunization, ii) they had received a boost of either Sputnik V or Moderna within 18 weeks post-prime dose, and iii) they presented to monitor their humoral immune response between 3-7 weeks after the boost dose. Individuals who received Sputnik V as the boost dose represented the homologous (SpV/SpV) group, while those who received the Moderna boost were included in the heterologous (SpV/Mod) group. The vaccination scheme depended on dose availability and the prioritization of risk populations, as established by the Argentine Ministry of Health at the time of boost.

### Immunogenicity

Binding IgG antibodies against the spike (S) receptor-binding domain (RBD) of SARS-CoV-2 (anti-S-RBD) concentration was assessed at 3-7 weeks after boost. Anti-S-RBD antibodies were quantified using the Abbott Diagnostics SARS-CoV-2 IgG II Quant chemiluminescent microparticle immunoassay (CMIA) on an Architect i2000 SR and an Alinity I analyzer (Abbott Diagnostics, Abbott Park, Illinois, USA). To standardize the results to WHO binding antibody units (BAU), a correction factor for Abbott arbitrary units (AU) was applied and where 1 BAU/mL=0.142 AU, as previously established by Abbott with the WHO international standard NIBSC 20–136^16^. Following the manufacturer recommendations, samples were considered as reactive for anti-S-RBD when titers were above 50 AU/mL (7.2 BAU/mL). An 80% protective effect (PROT-80) against symptomatic SARS-CoV-2 infection was assumed when anti-S-RBD titers were ≥506 BAU/ml^15^.

### Reactogenicity

All participants were invited to complete an online questionnaire to report all possible post-boost vaccination adverse events, medication, and medical assistance required. The intensity of adverse effects was graded as mild, moderate, and severe depending on the interference with daily activities.

### Statistical analysis

Descriptive statistics and univariate analyses were performed to compare the study groups. The outcome variable was the anti-S-RBD titer 3-7 weeks after the boost dose. Differences in anti-S-RBD levels and PROT-80 between the SpV/SpV and the SpV/Mod prime-boost schemes were evaluated. Categorical variables were expressed as number (percentage) and analyzed using the Chi-square test or the Fisher’s test while the student’s t-test and the Mann-Whitney *U* test were used for comparing continuous variables, which were expressed as median (interquartile range, IQR). Associations between anti-S-RBD levels and the time intervals from prime to boost (ΔP-B) and boost to anti-S-RBD IgG serological determination (ΔB-antiSRBD), as well as the age, were evaluated by means of the Spearman correlation coefficient (ρ). Those factors potentially associated with the outcome variable, such as age and sex, were evaluated in a generalized linear model. Finally, multivariate logistic regression models were developed to identify factors associated with PROT-80. Statistical analyses were carried out using the SPSS statistical software package release 23.0 (IBM SPSS Inc., Chicago, IL, USA).

### Ethical aspects

The study was designed and performed according to the Helsinki declaration and all blood donors gave their written informed consent (Study protocol EX-2021-06438339--UBA-DME#SSA_FFYB, Ethics committee of the Facultad de Farmacia y Bioquímica, Universidad de Buenos Aires).

## RESULTS

### Study population

From December 2020 to August 2021, 190 subjects were included in the study, 105 in the SpV/SpV group and 85 in the SpV/Mod group. Female participants were 132 (69.5%) and the median (IQR) age was 54 (37-63) years. Overall, median time intervals were 33 (24-97) days for ΔP-B and 23 (21-32) days for ΔB-antiSRBD, respectively. Forty-six (24.2%) individuals had a confirmed SARS-CoV-2 infection prior to vaccination, with a median (IQR) time of 21 (7.3-27) weeks between infection and prime dose as available data from 44 persons. Thus, 144 (75.8%) were *naïve* to SARS-CoV-2 infection at vaccination time. Table 1 shows epidemiological and vaccination-specific characteristics according to the vaccination group.

**Table 1.** Population epidemiological characteristics and vaccination-specific parameters.

| Characteristic | Homologous<br>SpV/SpV (n=105) | Heterologous<br>SpV/Mod (n=85) | p |
| --- | --- | --- | --- |
| Age* (years) | 42 (33-54) | 63 (56-66) | <0.001 |
| Female gender n (%) | 80 (76.2) | 52 (61.2) | 0.025 |
| Prior confirmed COVID-19 n (%) | 36 (34.3) | 10 (11.8) | <0.001 |
| $\Delta P$-B (days)* | 25 (21-27) | 97 (89-109) | <0.001 |
| $\Delta B$-antiSRBD (days)* | 26 (21-39) | 22 (21-26) | 0.001 |

SpV = Sputnik V, Mod = Moderna
\*Median (interquartile range)
$\Delta P$ -B: time from prime to boost; $\Delta B$ -antiSRBD: time from boost to anti-S-RBD IgG serological determination

### Immunogenicity

Anti-S-RBD IgG was reactive in all 190 (100%) samples. In the overall population, median (IQR) anti-S-RBD titers were 582 (209-1609) BAU/mL in individuals who received the SpV/SpV scheme, and 2511 (1476-3992) BAU/mL in the SpV/Mod group, p<0.001. Among women, anti-S-RBD was 1377 (347-2639) BAU/mL as compared to 1343 (503-3513) BAU/mL in men (p=0.438). In the bivariate correlation analyses among those who received the SpV/SpV or the SpV/Mod vaccination scheme, there was no significant association between anti-S-RBD levels and ΔP-B [ρ=0.033 (p=0.739) and -0.154 (p=0.160)], ΔB-antiSRBD [ρ=-0.128 (p=0.192) and -0.003 (p=0.981)] or age [ρ=-0.128 (p=0.192) and -0.003 (p=0.981)], respectively. Anti-S-RBD levels according to epidemiological and vaccination-specific parameters for the homologous and heterologous schemes are shown in Table 2.

**Table 2.**
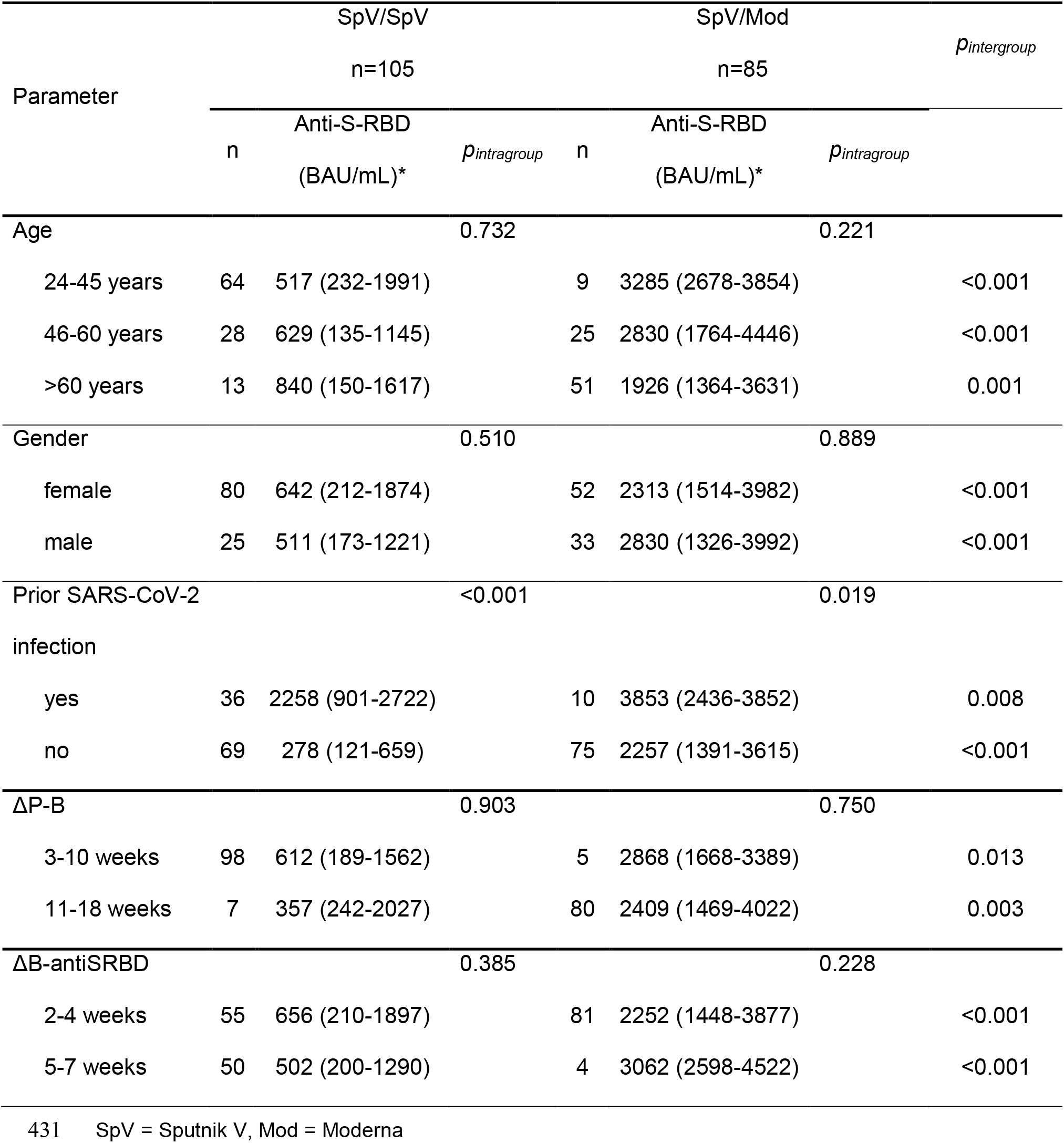
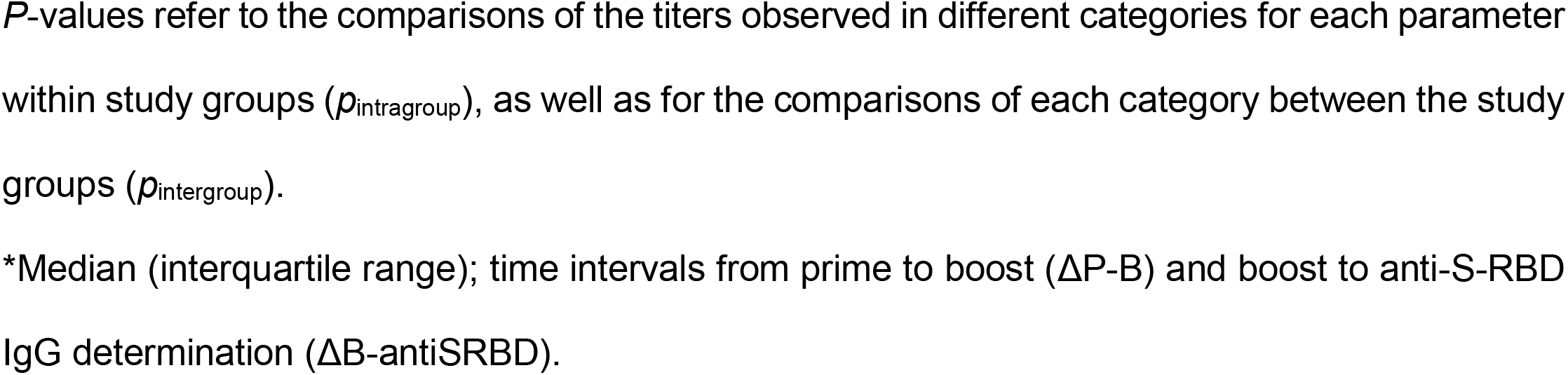
Anti-S-RBD IgG titer for homologous and heterologous vaccination schemes.

In participants of the SpV/Mod group who had a confirmed SARS-CoV-2 infection prior to vaccination, the humoral response as measured by anti-S-RBD levels was 1.7-fold higher than that observed in the non-infected ones. Likewise, within the SpV/SpV group, a confirmed previous SARS-CoV-2 infection resulted in 8-fold higher anti-S-RBD levels as compared to participants without prior infection (Table 2). Anti-S-RBD (IQR) levels were 2375 (722-4057) BAU/mL in subjects with a time difference between SARS-CoV-2 infection to prime dose above the median (21 weeks) as compared to 2618 (1712-4027) BAU/mL in patients who had suffered infection less than 21 weeks prior to prime vaccination (p=0.372). Achievement of PROT-80 were 20 (91%) versus 22 (100%), p=0.488, respectively.

Figure 1 shows the comparison of anti-S-RBD levels observed in SpV/SpV and SpV/Mod groups according to whether the participants had a confirmed SARS-CoV-2 infection prior to vaccination or not. The SpV/Mod vaccination scheme [B:2200; 95% confidence interval (CI):568-3832; p=0.009], as well as prior SARS-CoV-2 infection (B:1603; 95CI:1154-2051; p<0.001), were independently associated with anti-S-RBD levels in a generalized linear model adjusted for age (B:-1082; 95%CI:-17255-15089; p=0.895), gender (B:109076; 95%CI:-301717-519870), ΔP-B (B:-14323; 95%CI:-42977-14331; p=0.325) and ΔB-antiSRBD (B:-32095; 95%CI:-71911-7720; p=0.113).

**Figure 1.**
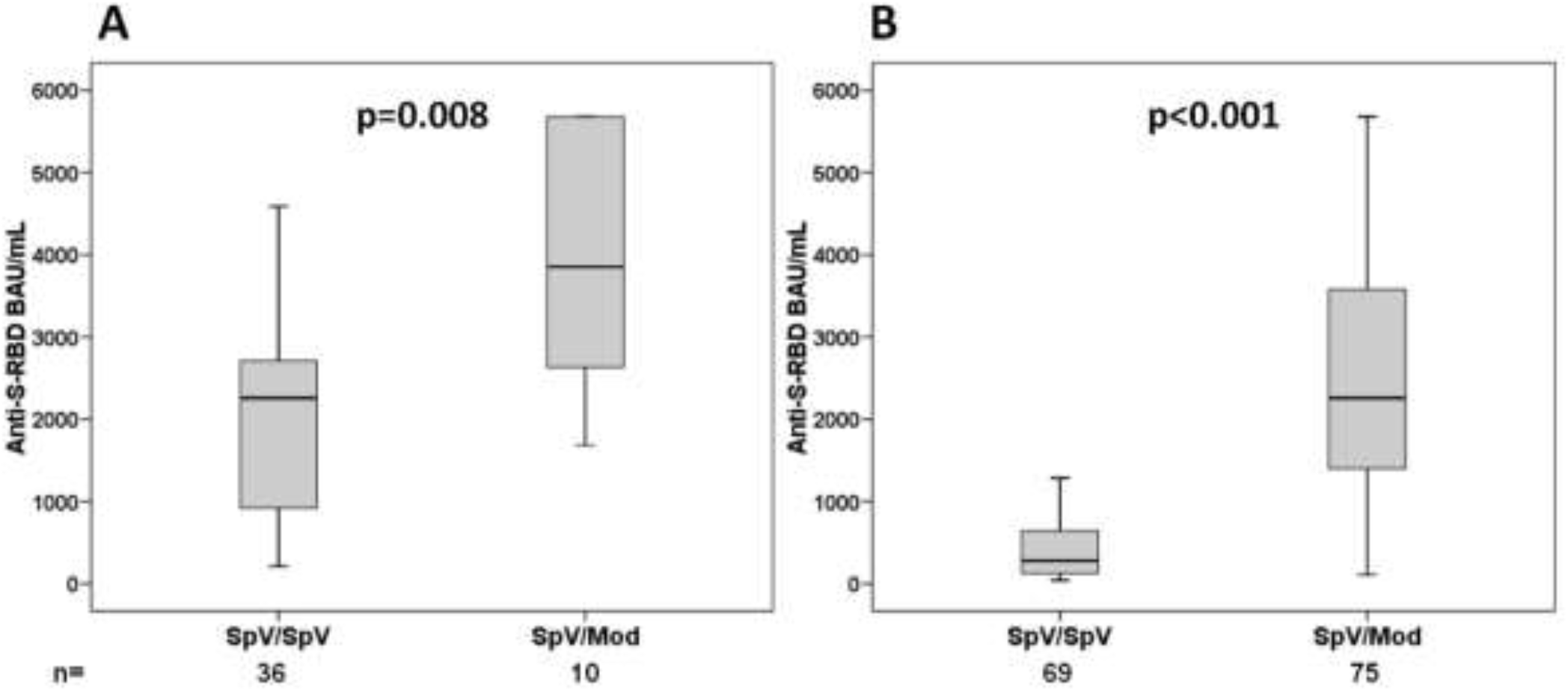
Anti-S-RBD IgG levels for homologous (SpV/SpV) and heterologous (SpV/Mod) scheme vaccinated subjects who had (A) or had not (B) a confirmed SARS-CoV-2 infection prior to vaccination.

The PROT-80 overall rate was 73.2%, as reached by 139 participants: 57 (54.3%) of the SpV/SpV group and 82 (96.5%) of the SpV/Mod group, p<0.001. The PROT-80 rates according to age, sex, prior SARS-CoV-2 infection, ΔP-B, and ΔB-antiSRBD are displayed in Table 3. Multivariate analyses identified the SpV/Mod scheme and a previous infection with SARS-CoV-2 as independent predictors to reach PROT-80 (Table 3).

**Table 3.** Univariate and multivariate analyses of potential predictors of the achievement of 80% protection against CoVID-19 (PROT-80) (n=190).

| Parameter | PROT-80, n (%) | | | $p_{uv(overall)}$ | Adjusted OR | $p_{mv}$ |
| --- | --- | --- | --- | --- | --- | --- |
|  | SpV/SpV | SpV/Mod | Overall |  | (95% CI) |  |
| Vaccine scheme |  |  |  |  |  |  |
| SpV/SpV |  |  | 57 (54.3) | <0.001 | Ref | <0.001 |
| SpV/Mod | - | - | 82 (96.5) |  | 4.154<br>(6.585-615.55) |  |
| Age (years)* |  |  |  |  |  |  |
| 24-45 | 35 (54.7) | 9 (100) | 44 (60.3) | 0.003 | -0.007 | 0.993 |
| 46-60 | 15 (53.6) | 25 (100) | 40 (75.5) |  | (0.957-1.029) |  |
| >60 | 7 (53.8) | 48 (94.1) | 55 (85.9) |  |  |  |
| Gender |  |  |  |  |  |  |
| Male | 13 (52) | 31 (9.9) | 44 (75.9) | 0.577 | Ref | 0.686 |
| female | 44 (55) | 51 (98.1) | 95 (72.0) |  | 0.436<br>(0.545-4.387) |  |
| Prior SARS-CoV-2 infection |  |  |  |  |  |  |
| No | 23 (33.3) | 72 (96) | 95 (66.0) | <0.001 | Ref | <0.001 |
| Yes | 34 (94.4) | 10 (100) | 44 (95.7) |  | 3.732<br>(8.641-202.01) |  |

| $\Delta P$-B (weeks) | | | | | | |
| --- | --- | --- | --- | --- | --- | --- |
| 11-18 | 3 (42.9) | 77 (96.3) | 81 (92.0) | <0.001 | Ref | 0.754 |
| 3-10 | 54 (55.1) | 5 (100) | 58 (56.9) |  | -0.004 |  |
|  |  |  |  |  | (0.972-1.021) |  |
| $\Delta B$-antiSRBD (weeks) | | | | | | |
| 3-4 | 32 (58.2) | 72 (96) | 104 (80.0) | 0.002 | Ref | 0.125 |
| 5-7 | 25 (/50) | 10 (100) | 35 (58.3) |  | -0.036 |  |
|  |  |  |  |  | (0.921-1.01) |  |

SpV: Sputnik V; Mod: Moderna; uv: univariate; mv: multivariate; P: prime; B: boost. *P*-values for age, gender, prior SARS-CoV-2 infection, $\Delta P$ -B and $\Delta B$ -antiSRBD were 0.955, 0.486, <0.001, 0.404 and 0.26 for the SpV/SpV group and 0.355, 0.333, 0.684, 0.832 and 0.684 for the SpV/Mod group, respectively. \*Entered as continuous variable in the multivariate analysis.

### Reactogenicity

The analysis of the reactogenicity was evaluated in both groups during seven days after the boost. The homologous and heterologous immunization schemes were well tolerated, and no medical assistance or potentially life-threating events were reported. Adverse events, including local and systemic symptoms, were more frequent in the SpV/Mod group (72.9%) than in the SpV/SpV group (55.2%), p=0.012. Overall, the most frequent systemic adverse events were myalgia (26.8%), fever (21.6%), fatigue (19.5%), and headache (16.8%). The heterologous vaccine scheme induced significantly more systemic adverse effects than the homologous one (64.7% vs 36.2%, p<0.001). Table 4 shows the presence and intensity of systemic adverse effects by vaccination scheme. Regarding local adverse events, pain at the injection site was frequently reported (20.0%) and it was significantly more frequent for the homologous scheme than for the heterologous one (28.0% vs 11.8%, p=0.011). Figure 2 shows the reactogenicity by adverse effect (local and systemic) according to the vaccination scheme.

**Table 4.** Presence and intensity of adverse systemic effects by vaccination scheme.

| **Intensity**<br>**of adverse event** | **Homologous**<br>**SpV/SpV (n=105)** | **Heterologous**<br>**SpV/Mod (n=85)** | **p** |
| --- | --- | --- | --- |
| No reaction | 67 (63.8%) | 30 (35.3%) | <0.001 |
| Mild | 13 (12.4%) | 20 (23.5%) | 0.043 |
| Moderate | 14 (13.3%) | 27 (31.8%) | 0.002 |
| Severe | 11 (10.5%) | 8 (9.4%) | 0.807 |

SpV = Sputnik V, Mod = Moderna

**Figure 2.**
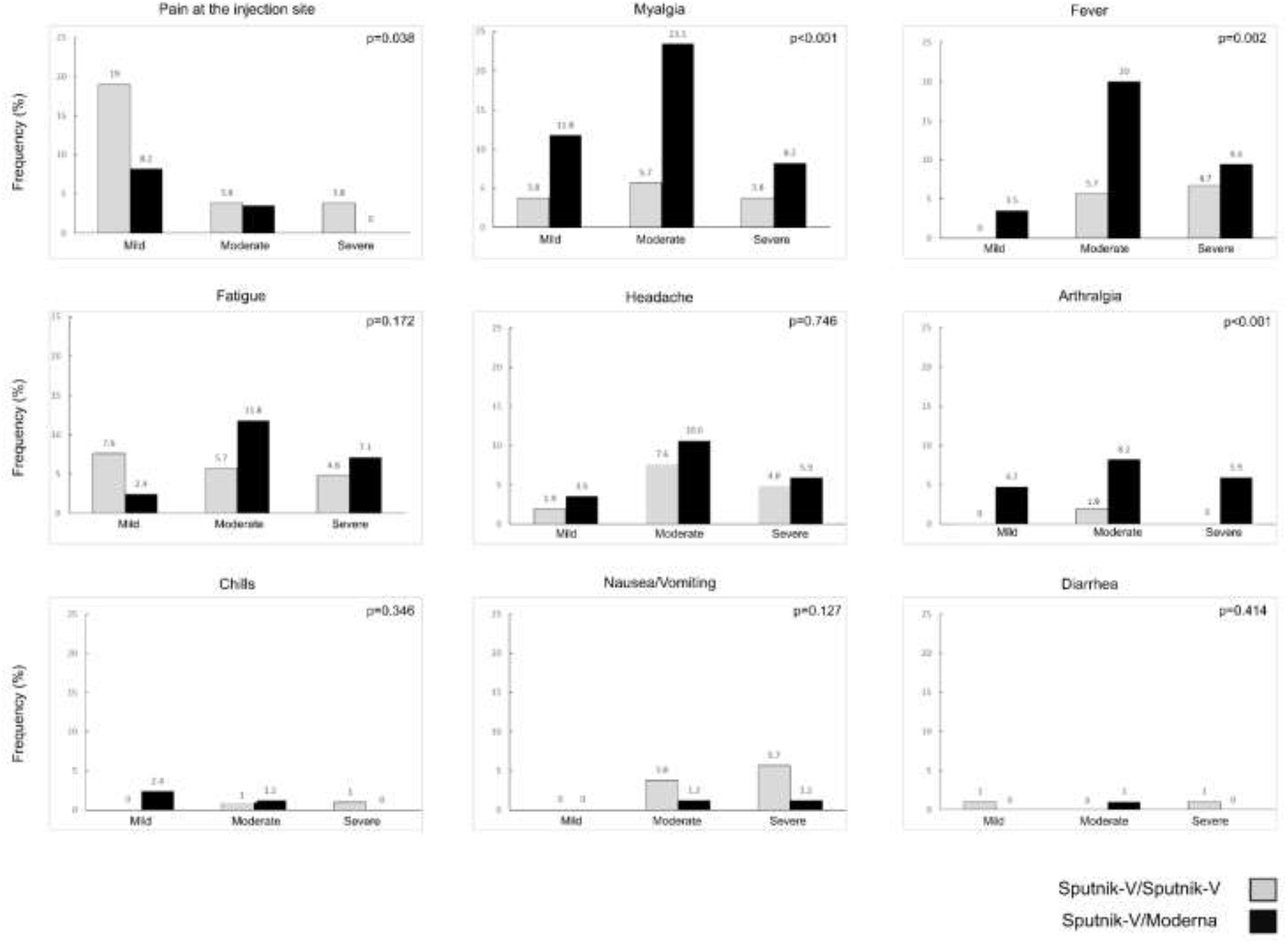
Reactogenicity: frequency of participants’ reported local and systemic adverse effects for the SpV/SpV and SpV/Mod schemes, classified by severity

## DISCUSSION

The present work shows that, in a real-life setting, the incorporation of heterologous prime-boost vaccination against SARS-CoV-2 could be a valuable strategy in response to deficiencies in doses supply or the presence of serious adverse effects in the prime dose. Overall, the heterologous scheme showed significantly higher titers of anti-S-RBD IgG. Moreover, both vaccination schemes were well tolerated, and no medical assistance was required.

All the study participants presented anti-S-RBD IgG seroconversion after the complete immunization with both boosts. These results were expected for the Sputnik V complete scheme^13,17^ and validated the immunogenicity achieved with the Moderna boost. Importantly, a higher antibody titer was achieved with the heterologous scheme. Although a better humoral response has been already described for mRNA vaccines as compared to adenovirus-based^3,4,7,8,18,^ there is still no data validating this assumption for the SpV/Mod combination. Our results support the use of the Moderna vaccine as an alternative to the homologous Sputnik V scheme in general. Also, this would be of special interest during a supply shortage. Further cellular studies are justified since the combination of vaccines appears to complement the characteristic immune response triggered by each vaccine platform^7,19^. In this way, the SpV/Mod combination could especially benefit certain risk groups through the development of a more robust and long-lasting immune response against SARS-CoV-2.

Although the threshold to confer protection against SARS-CoV-2 after COVID-19 vaccination depends on several factors, a randomized trial on the ChAdOx1 vaccine efficacy provides robust data to deduct the antibody levels associated with protection against SARS-CoV-2. In this trial, Feng et al identified 506 BAU/ml as the cut-off level for binding anti-S-RBD antibodies to provide a vaccine efficacy of 80% against symptomatic SARS-CoV-2 infection^15^. According to this threshold, in the present study, almost 100% of the SpV/Mod group had a vaccine efficacy of 80% while only 54% of the SpV/SpV group achieved such protection. Moreover, in the multivariate analysis, the vaccine efficacy of 80% was not associated with age, gender, ΔP-B, and ΔB-antiSRBD. All this is in accordance with previous studies on homologous regimens that, when compared, suggest that Sputnik V is moderately less effective than Moderna vaccine in preventing symptomatic COVID-19^13,18,20^.

In the present study, no significant gender-specific differences in anti-S-RBD IgG titers were observed. This result is in line with other studies where gender seems not to influence antibody development against Sputnik V and Moderna vaccines^10,17,21,22^. In contrast, other studies have reported higher antibody levels in women^23-24,^ reflecting the need for further research in this field. Likewise, contradictory results have been published related to the age of the vaccinees. While some studies have reported higher antibody titers in young people^24-26^, Wheeler and colleagues did not find an antibody response-age dependency after the second dose of the Moderna vaccine^22^. In another study carried out in Argentina, involving a population with a mean age of 45 years, Rovere et al. also failed to detect an age-response correlation for the Sputnik V scheme^17^. In accordance with the latter findings, we did not observed differences in the anti-S-RBD IgG titers related to participants’ age.

As expected, a significant correlation was found between higher anti-S-RBD IgG titers and a confirmed prior infection with SARS-CoV-2 for both schemes. However, the impact of time between infection and prime dose did not reach statistical significance. This finding is in line with previous studies reporting an increased anti-S-RBD IgG response in subjects previously infected with SARS-CoV-2^10,17,27^. Importantly, in the present setting, the difference between schemes was more evident for the *naïve* individuals, reaching 8-fold higher anti-S-RBD IgG titers for the heterologous scheme with respect to the homologous one. It can thus be concluded that especially those without prior infection would benefit from a switch in the boost component.

Regarding reactogenicity, adverse events including local and systemic symptoms in participants who received SpV/Mod vaccination was slightly higher than for SpV/SpV, confirming previously reported findings^13,21.^ Importantly, these findings are derived mainly from participants of a clinical trial and there is very little data on SpV/SpV reactogenicity under real-life conditions. The present work therefore provides important contribution to the understanding of vaccination in the clinical practice. In contrast, the higher rate of adverse events in the SpV/Mod scheme was expected since it is well established that stronger side effects are associated with mRNA platforms^20,28,29.^ However, it is important to note that very similar severe adverse events were observed (10% SpV/Mod vs 10.5% SpV/SpV) and no major effects requiring medical assistance were reported in any of the schemes. Moreover, it should be considered that, since most serious adverse effects have been reported in a very low frequency, even when no participant required medical assistance, the small size of our sample could have influenced on this aspect^30^.

This study has some limitations. First, due to the observational, real-world design of our study, the vaccination scheme distribution was not randomized and depended on the risk priorities established by the Ministry of Health. Due to the limited availability of doses, the SpV/SpV group was mostly formed by active healthcare-workers who were at highest priority for vaccination at the time and benefited from the last available Sputnik V boost doses. In contrast, the SpV/Mod scheme was mostly applied to individuals older than 60 years old who were at intermediate priority. Apart from age distribution, the limited supply of the second component of Sputnik V led to almost 4-fold differences in the median interval between prime and boost immunization, since the Moderna vaccine was also not immediately available once the shortage of Sputnik V was evident. However, there was no correlation between age or the time intervals used in this study and the anti-S-RBD levels. As a result, when the samples were stratified according to age, ΔP-B, and ΔB-antiSRBD, no significant differences of the antibody titers were observed between different levels of categorization within each vaccine scheme group, while the significant difference in anti-S-RBD between the two vaccine scheme groups was maintained. Therefore, both populations can be regarded as comparable and an impact of differences in age and time intervals on the analyses is very unlikely in the present setting. Second, due to the lack of availability in Argentina, a homologous Moderna vaccination control group could not be included to determine whether the elevated anti-S-RBD levels were due to an additive or a synergistic effect of the two vaccines. However, data from other studies on Mod/Mod suggest a strong immune response to anti-S-RBD^22^. Still, future studies are warranted to clarify this issue.

In conclusion, the heterologous immunization with SpV/Mod is not only immunogenic and well tolerated but also induces a stronger humoral response than the homologous Sputnik V scheme. Further studies such as, the neutralization plaque assay against the current and emerging variants of SARS-CoV-2, are needed to investigate the efficacy of SpV/Mod vaccinations in the local realities of each population.

## Data Availability

All data produced in the present work are contained in the manuscript

## ACKNOWLEDGMENT

Matías J. Pereson, Patricia Bare, María Noel Badano and Federico A. Di Lello are members of the National Research Council (CONICET) Research Career Program. Karin Neukam is the recipient of a Miguel Servet contract by the Instituto de Salud Carlos III (grant number CPII18/00033). We would like to thank to Mrs. Silvina Heisecke, from CEMIC -CONICET, for the copyediting of the manuscript

## Authorship

Matías J. PERESON and Karin NEUKAM: Analysis and interpretation of data, Drafting the article. Final approval of the version to be submitted.

Lucas AMAYA: Acquisition, analysis, and interpretation of data. Final approval of the version to be submitted.

Patricia BARE, Natalia ECHEGOYEN, María Noel BADANO, Alicia LUCERO, Antonella MARTELLI: Acquisition of data, Final approval of the version to be submitted. Gabriel H. GARCIA, Cristina VIDELA, Alfredo P. MARTíNEZ: Analysis and interpretation of data, Revising the manuscript critically for important intellectual content, Final approval of the version to be submitted.

Federico A. DI LELLO: Conception and design of the study, Acquisition, Analysis, and interpretation of data, Drafting the article

